# Leveraging global PhPID framework to enable more granular signal detection and characterization in VigiBase: a dexamethasone case study

**DOI:** 10.64898/2026.07.13.26357959

**Authors:** Paula Vasconcelos-Blomberg, Joana Félix China, Bushra Riaz Syeda, Malin Fladvad, Olof Lagerlund, Lucie M. Gattepaille, Michele Fusaroli

## Abstract

**Introduction:** Conventional substance-level disproportionality analysis may miss safety patterns specific to a dose form, route, or intended site. More granular analyses are hindered by incomplete, inconsistent reporting of product information. The Pharmaceutical Product Identifier (PhPID), representing products by substance, strength, and dose form, may support more granular analyses.

**Objective:** To explore the use of PhPID-like dose form information for site-specific disproportionality analysis in dexamethasone.

**Methods:** We evaluated VigiBase reports (January 1, 2001 - December 31, 2024) for completeness of dose form and route data. We standardized dexamethasone entries to PhPID Level 3 standards, representing substance and administrable dose form. Through disproportionality analysis (Information Component, IC) we compared substance-level and site-specific results.

**Results:** Among 56.4 million suspected/interacting drugs, dose form was reported in 47.7%, route in 69.4%. Among 109,248 dexamethasone entries, 703 dose form and 80 route variations were mapped to 53 and 44 standard codes respectively; about half could be mapped unambiguously. Site-specific analyses revealed biologically plausible patterns not apparent in substance-level analyses. Ocular use showed higher ICs for glaucoma and cataract, while systemic use showed higher IC for psychiatric and endocrine events (e.g., depression, agitation, Cushing’s syndrome). IC time-trends suggested that some signals (e.g., cataract with Ocular use) could emerge earlier in site-specific analyses.

**Conclusion:** More granular product information, aligned with PhPID, may improve signal detection and characterization of site-specific safety issues. These findings support granular identifiers in pharmacovigilance while highlighting the need for better capture and standardization of dose form and route of administration data.

**Key Points:**

- Conventional substance-level disproportionality analysis can miss administration site-specific safety patterns, but incomplete and heterogeneous reporting of dose form and route of administration remains a major barrier to more granular analyses in VigiBase.
- The Pharmaceutical Product Identifier (PhPID) provides a framework for representing medicinal products with higher granularity than the active substance alone, using dose form and other product attributes to support more granular pharmacovigilance analyses.
- When standardization is available, site-specific disproportionality can reveal biologically coherent safety patterns that may be obscured in aggregated substance-level analyses.

## 1. Introduction

Drugs are available in multiple pharmaceutical forms (e.g., oral tablets, injectable solutions, topical creams) and routes of administration (e.g. oral, parenteral, transdermal). As a result, products containing the same active ingredient can differ in pharmacokinetics and distribution. Such a difference may influence the type, frequency, and severity of adverse drug reactions (1,2) and, in some cases, lead to distinct overall safety profiles (3,4). Corticosteroids (e.g., dexamethasone) illustrate this well: they are marketed in heterogeneous formulations and for several administration sites, and, for example, periocular and intraocular routes cause elevated intraocular pressure more often than oral or cutaneous routes (5). Route- and formulation-dependent differences in safety profiles have also been described for semaglutide (6) and 5-aminosalicylate (7).

Case by case review can hint to formulation-specific safety signals (8). Instead, disproportionality analysis, a necessary complementary prioritizing tool in settings with high volumes of reports (9), cannot differentiate by formulation or route of administration in its traditional implementation at the substance (active ingredient) level. Consequently, formulation-specific safety issues may remain undetected, be detected late, or be attributed too broadly to the substance irrespective of formulation.

A major barrier to more granular pharmacovigilance analyses is the incomplete and inconsistent capture of medicinal product information in adverse event reports (10). The ICH (International Council for Harmonisation of Technical Requirements for Pharmaceuticals for Human Use) E2B standard for electronic transmission of reports includes fields for product name, substance, dose form, strength, and route of administration^1^(11,12). In practice, however, drug information is often limited to free text describing the product or substance, while more granular fields are left empty or recorded using heterogeneous terminologies^2^. In VigiBase–the WHO global database of adverse event reports for medicines and vaccines(15)–free text drug names are standardized to substances using the Uppsala Monitoring Centre’s (UMC’s) Drug Dictionary WHODrug Global–a global medicine and vaccine terminology for identification of medicinal products (16)–, but no equivalent mapping has routinely been applied for additional attributes such as dose form, strength, and route of administration.

One opportunity for more comprehensive standardization comes from the International Organization for Standardization (ISO) Identification of Medicinal Products (IDMP) standards, first published in 2012 (17). Within this framework, the Pharmaceutical Product Identifier (PhPID; ISO 11616:2017), defines pharmaceutical products on the basis of active substance, strength, and administrable dose form (10,18). PhPID supports several levels of granularity, ranging from L1 (substance only) to L4 (substance, strength, and dose form) (Figure 1). As global implementation and integration of PhPIDs into adverse event reports progresses, evidence is needed on the practical value of this enhanced granularity for pharmacovigilance (19).

**Figure 1.**
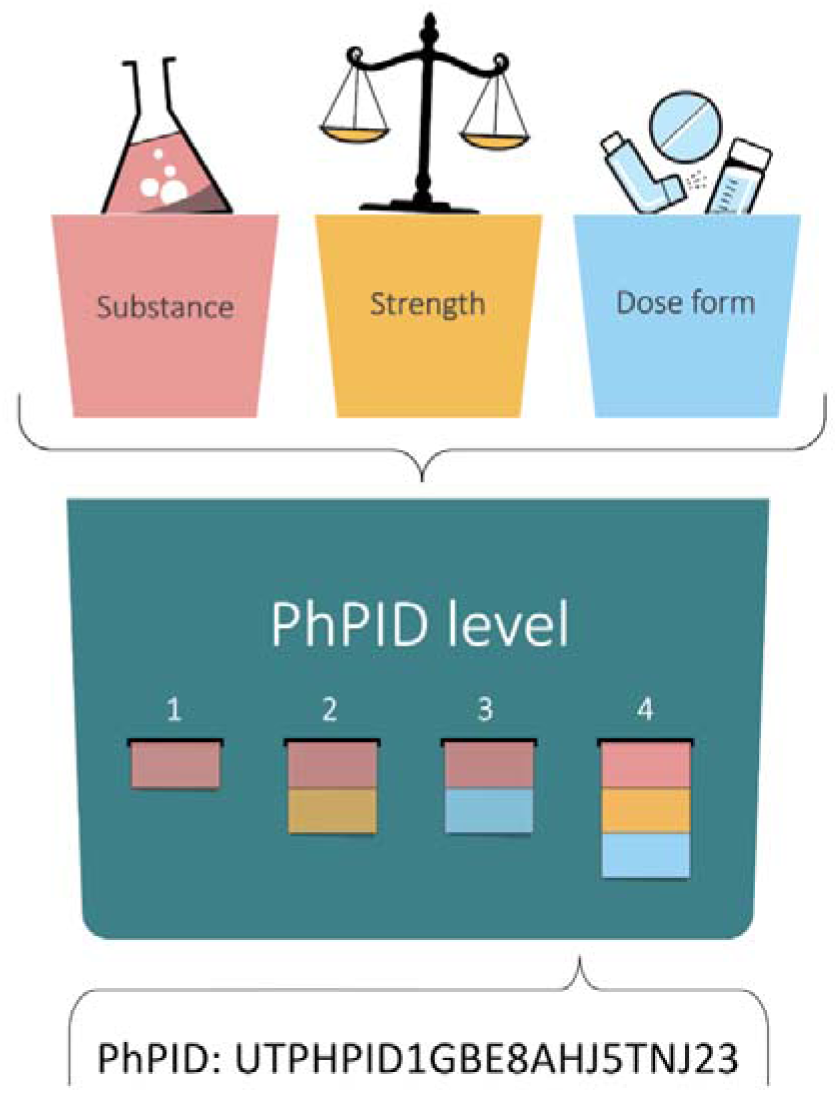
The four levels of Pharmaceutical Product Identifier (PhPID) from (18). Level 1 substance, level 2 substance and strength, level 3 substance and dose form and level 4 substance, strength and dose form, Note that the format of the PhPID is now changed.

**Figure 2.**
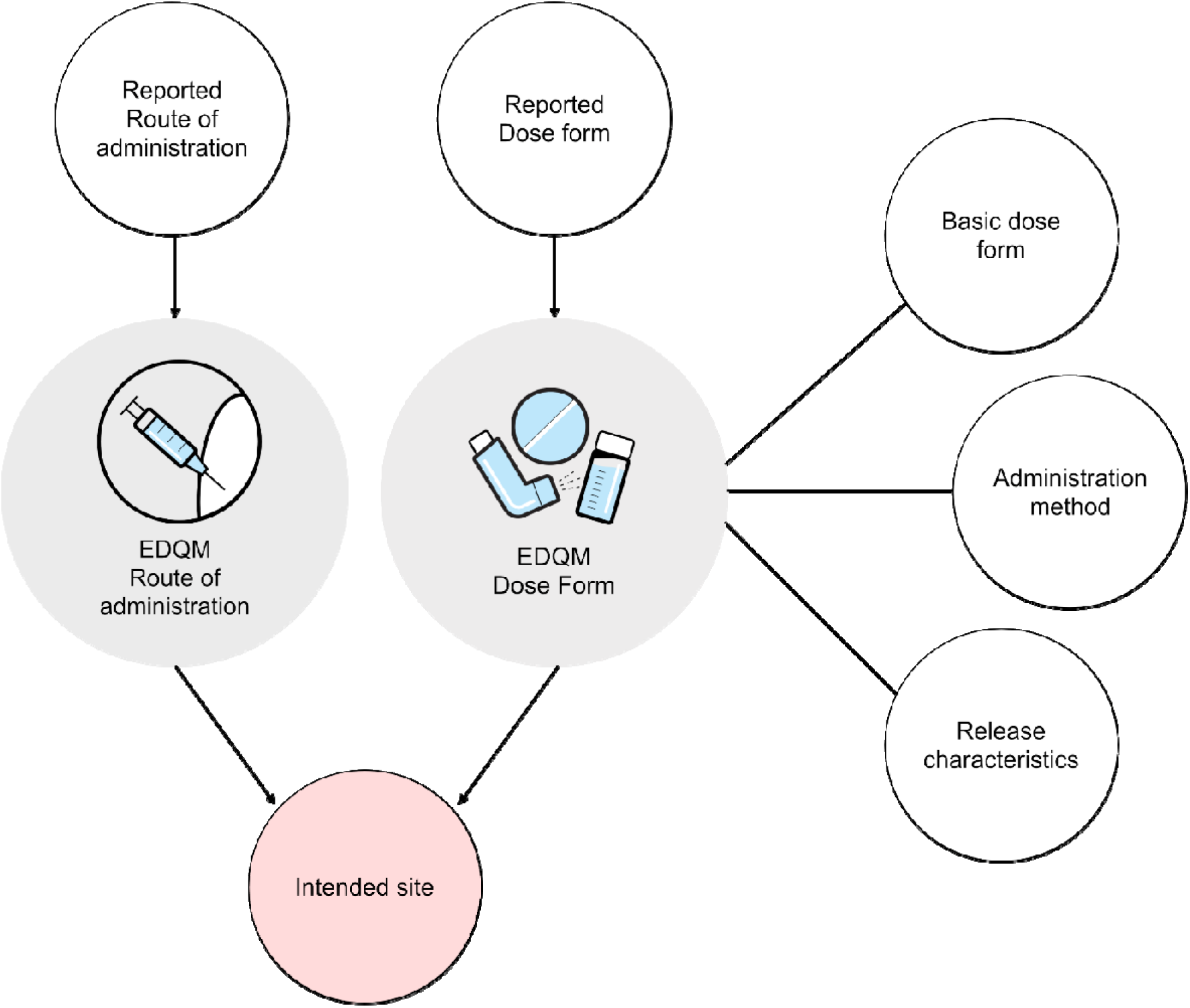
The stepwise mapping procedure to EDQM standardised terms according to (18)

At the time this study was designed and conducted, however, PhPIDs were not routinely available in adverse event reports, instead, we used a standardization approach based on the global PhPID framework and internationally available EDQM Standard Terms for pharmaceutical dose forms, routes of administration, and dose form attributes. This allowed us to use the EDQM dose form attribute intended site of administration as an approximation of the type of granularity envisioned by PhPID while avoiding premature dependence on a specific identifier implementation in an exploratory study.

In this study, we explore whether administration site-level identifiers can strengthen signal detection and characterization in VigiBase by enabling more granular identification of administration site-specific safety issues.

## 2. Methods

This study was conducted in VigiBase, the WHO global database of adverse event reports for medicines and vaccines, containing over 40 million reports submitted by members of the WHO Programme for International Drug Monitoring (15). We focused on PhPID Level 3, namely Substance and Administrable Dose Form, with particular attention to the dose-form attributes administration site (Figure 1).

Because route of administration and dose form in VigiBase are not yet harmonized to a single standard terminology, we therefore explored the feasibility and utility of an approach based on the PhPID framework where we standardized route and dose form information. Full manual standardization across all substances in VigiBase was not feasible, so we selected dexamethasone as a use case. Dexamethasone was selected because it combines broad use, diverse dose forms and routes of administration, sufficient reporting volume, and known biologically plausible intended site-dependent adverse reactions, making it a suitable proof-of-concept case for administration site-specific analyses. The study proceeded in three steps: (i) characterization of the availability of route and dose form information in VigiBase, (ii) manual standardization of dexamethasone route and dose form data to EDQM terminology to construct administration site-level identifiers (13), and (iii) administration site-aware disproportionality analyses for selected adverse events.

### 2.1 Characterization of available route and dose form information

To assess the availability of PhPID Level 3-relevant information in VigiBase, representing substance and administrable dose form, we extracted reports submitted from January 1, 2001, to December 31, 2024. This period was selected to capture reports in ICH E2B(R2) and later formats while minimizing inclusion of older reporting standards that would further complicate standardization. The analysis included drugs reported as either suspected or interacting. For each drug entry, we assessed whether dose form and route of administration were recorded and calculated annual completeness as the percentage of total drug entries with each field populated.

### 2.2 Manual mapping of route and dose form for dexamethasone

For the mapping, we included reports in ICH E2B(R2), E2B(R3), and INTDIS (International Drug Information System) where dexamethasone was listed as suspected or interacting. To characterize reporting heterogeneity, we identified all unique free-text entries for route of administration and dose form and tabulated their frequencies.

Original entries were then manually mapped by a medical doctor, and pharmacovigilance expert, when possible, to EDQM terminology (standards for pharmaceutical dose form and for route or method of administration (13)). Standardization was performed iteratively, using a custom mapping dictionary that combined official mappings from legacy standards to EDQM terminology (13,20,21), internally validated mappings for free-text entries, and multilingual variants. Route entries were mapped to EDQM Routes of Administration (RoA), and dose form entries to the closest corresponding EDQM dose form term (22). Standardized routes and dose forms were subsequently reviewed by a terminology expert. The administration site, as defined by EDQM dose form attributes, was assigned by following the PhPID business rules (Supplementary table), which define the intended site assignment for each combination of standardized route of administration and dosage form. Where a report’s exact route–dosage form combination was not covered by the rules, administration site was assigned using a fallback hierarchy: first by route alone (if the route was known but the specific dosage form was not covered), then by dosage form alone (if the route was missing or unresolvable). Reports for which no rule applied at any level were left unmapped and excluded from route-specific analyses. In this study, administration site was treated as a standardized proxy derived from reported route and dose form fields, and therefore approximated reported use (plausibly nearer to the actual site of administration) rather than directly capturing intended clinical site of administration.

We quantified completeness of dose form, route, and administration site before and after standardization. We also measured the reduction in the number of unique variants after harmonization (as multiple entries could be mapped to the same EDQM term) and assessed data loss resulting from unsuccessful mapping. This analysis was intended to illustrate both the efforts required to standardize these data in VigiBase and the effect of standardization on data retention.

### 2.3 Disproportionality Analysis

We selected a broad set of MedDRA Preferred Terms (PTs) corresponding to known reactions to dexamethasone which are considered likely to vary by administration site. The PT set was selected as a proof-of-concept panel to examine whether known or plausible administration site-dependent patterns could be recovered using a more granular exposure definition.

To assess the impact of incorporating administration site granularity into signal detection, we compared the Information Component (IC) with 95% credibility intervals (23) calculated using two exposure definitions. (1) In the substance-level reference analysis, exposure included all dexamethasone reports irrespective of administration site. (2) In the site-specific analysis, exposure was defined at the substance-site level and included only reports in which dexamethasone was reported for a given administration site (for example, Ocular use). Both analyses considered only suspected and interacting substances and used the crude database as a background to calculate the expected frequency. This background was chosen to preserve statistical power and to assess site-specific disproportionality against overall reporting in the database, rather than relative to a narrower subset of drugs sharing the same administration site.

The IC was computed as:

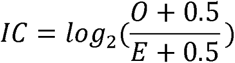

where *O* is the observed number of reports for the substance-event combination of interest (or the substance-site-event combination of interest for *O_site_*) and *E* is the expected count under the assumption of independence. The expected count was calculated for both analyses using the frequency of the event on the crude database:

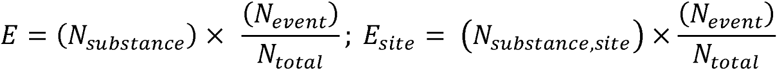

Where *N_substance_* is the total number of reports with dexamethasone (suspected or interacting), *N_substance,site_* is the same count but limited to the administration site of interest (e.g., dexamethasone-ocular), *N_events_*is the total number of reports with the event of interest, and *N_total_* is the total number of reports in the dataset.

To structure the interpretation of site-specific results, we formulated a priori expectations for the direction of site-specific disproportionality based on known pharmacology and the established safety profile of dexamethasone. Corticosteroid-induced ocular adverse reactions — particularly cataract, glaucoma, and elevated intraocular pressure — are well-documented consequences of local ocular exposure and were therefore expected to show stronger disproportionality for Ocular use than for systemic administration sites. Conversely, systemic adverse reactions attributable to hypothalamic-pituitary-adrenal axis suppression (e.g., Cushing’s syndrome, adrenal insufficiency), psychiatric effects (e.g., depression, mania, agitation, insomnia, psychotic disorder), and metabolic or musculoskeletal effects (e.g., diabetes mellitus, weight increased, osteoporosis, fracture) were expected to show stronger disproportionality for Oral and Parenteral use, given the higher systemic bioavailability of these routes. Gastrointestinal events such as peptic ulcer and gastrointestinal haemorrhage were expected to be more prominent for Oral use specifically, reflecting direct mucosal exposure.

## 3. Results

### 3.1 Characterization of available route and dose form information

Between January 1, 2001 and December 31, 2024, VigiBase collected 56.4 million drug entries coded as suspected or interacting. Dose form information was available for 47.7 % of entries, while route of administration was present in 69.4% (Figure 3). Completeness of route information declined over time from 90% in 2001 to 61% in 2024. Completeness of dose form information increased substantially after 2012 (13.9%), peaked in 2016 (68.8%), and then gradually declined down to 48.4% in 2024 (Figure 4).

**Figure 3.**
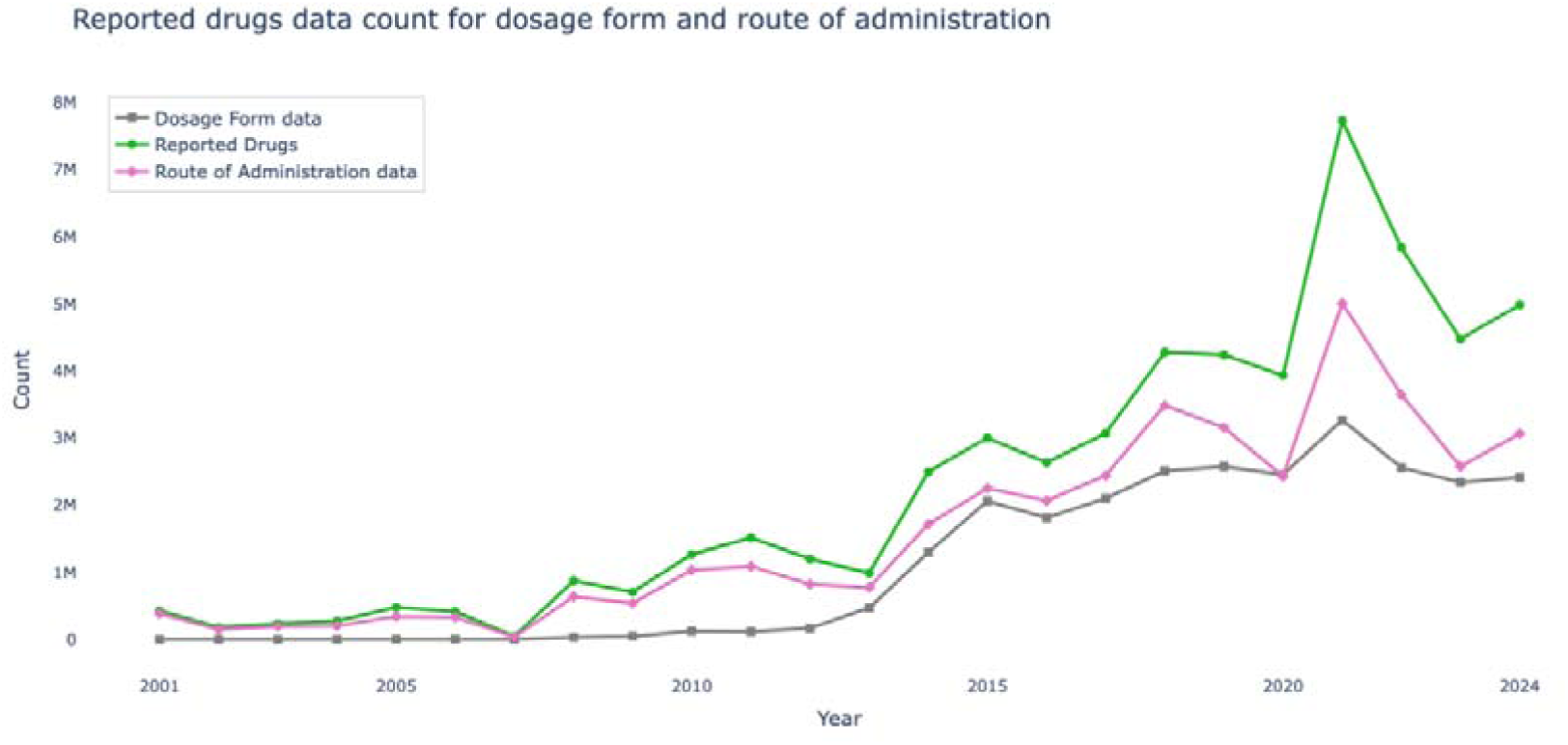
Annual counts of reported drugs in VigiBase (2001–2024), and corresponding counts of reported drugs with drug dosage form and route of administration data.

**Figure 4.**
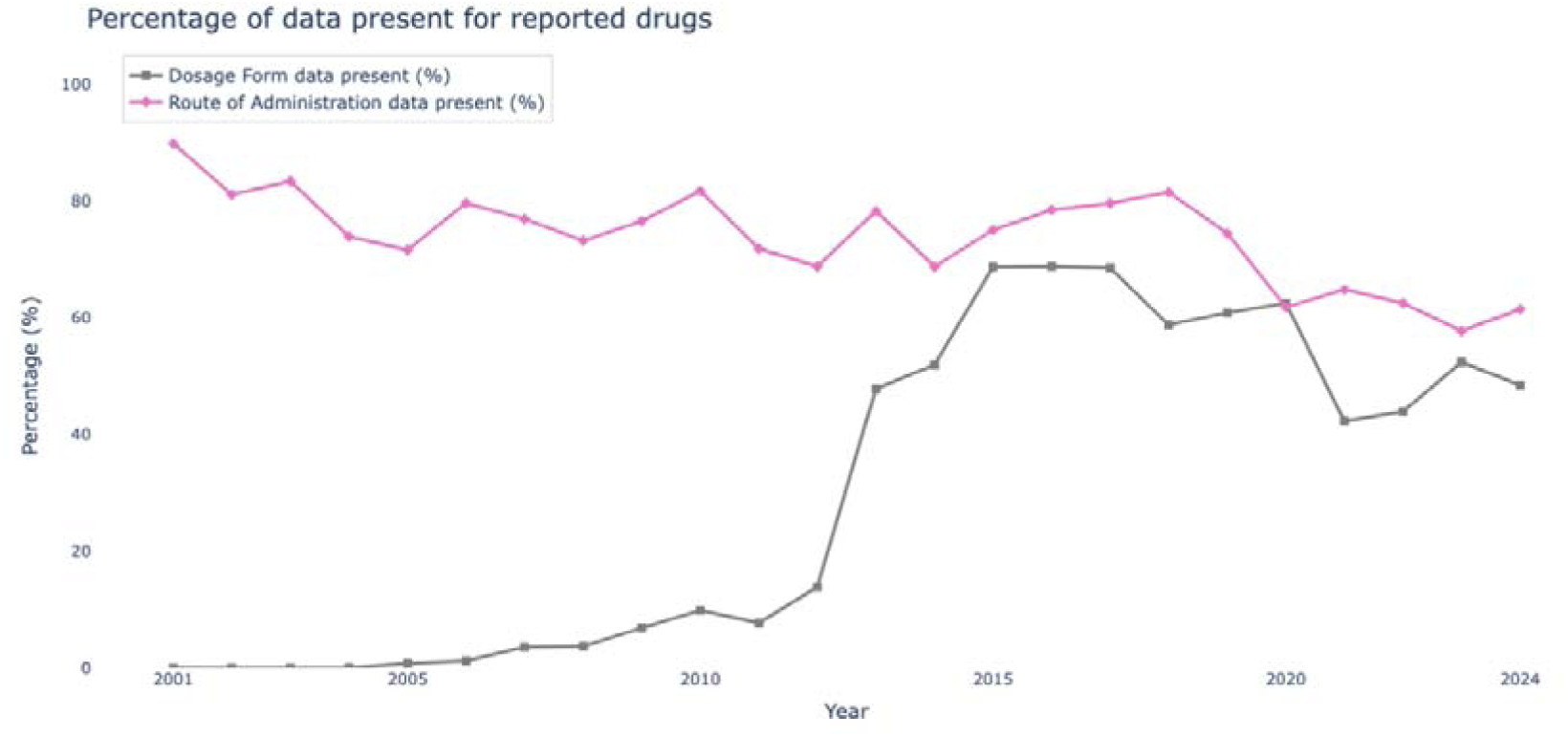
Percentage of reported drugs in VigiBase (2001–2024) that include dosage form and route of administration data.

### 3.2 Standardization and mapping of Route and Dose Form

We extracted 109,248 dexamethasone entries in ICH E2B(R2), E2B(R3), and INTDIS formats coded as suspected or interacting. For route of administration, 80 unique free text values were identified and mapped to 44 unique EDQM codes (PDF, PFT, BDF). Overall, 46.4% of entries were mapped, 27.4% were missing, and 26.2% could not be mapped because of ambiguity or insufficient detail (Figure 5). For dose form, 703 unique free text values were identified and mapped to 53 unique EDQM codes. Overall, 24.0% of entries were mapped, 59.9% were missing, and 16.1% could not be mapped.

**Figure 5.**
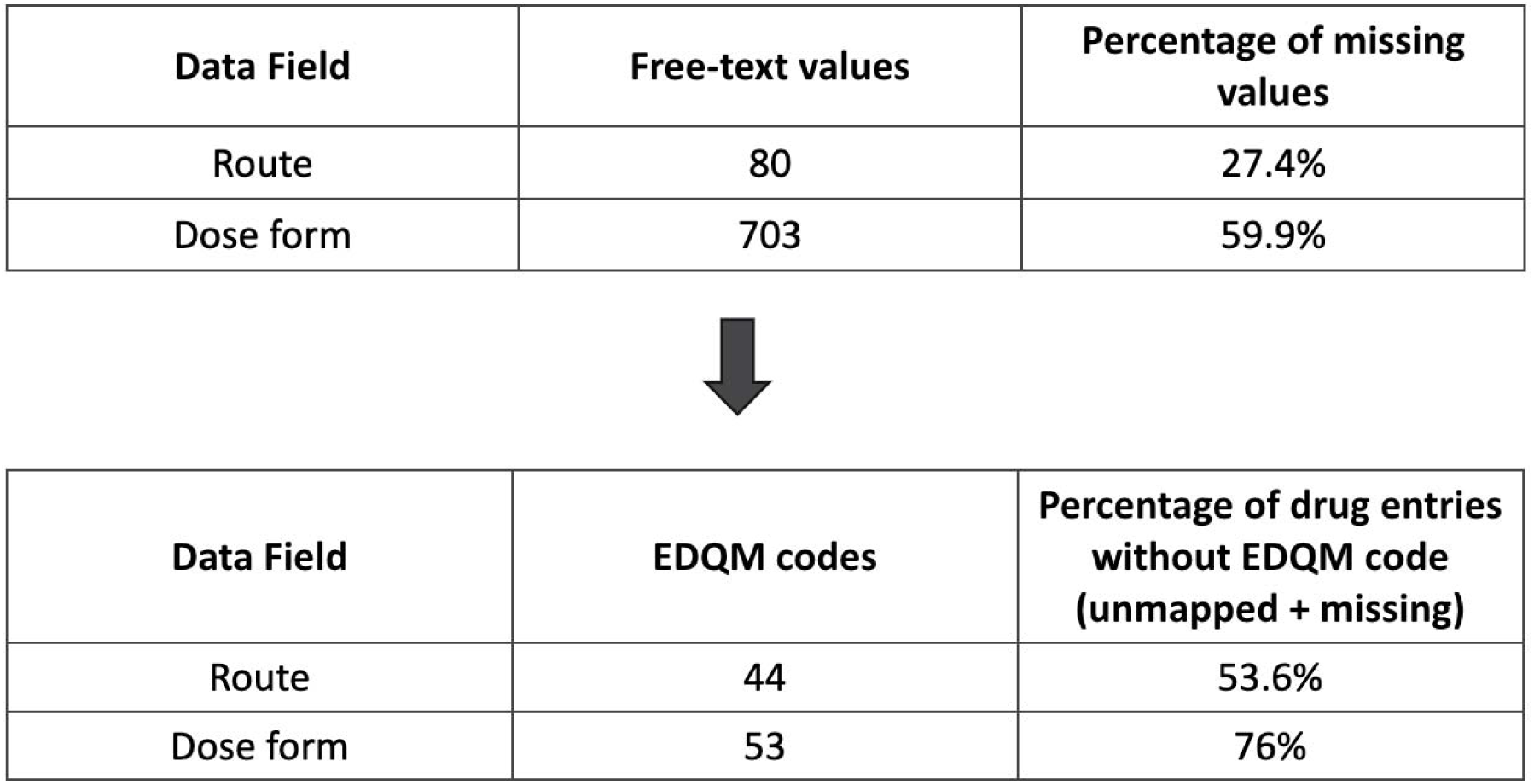
Changes in variability and mapping outcomes for route and dosage form data before and after standardization to the PhPID framework.

Taken together, approximately half of dexamethasone entries contained route or dose form information that could be standardized to EDQM terminology. Among standardized dose forms, Tablet was the most frequent (10%), followed by Solution for injection (5.4%) and Injection (5.0%), while Ophthalmic insert (1%) and Eye drops (0.3%) were rare (**Table 1**). Among standardized routes, Oral use (24.3%) and Intravenous use (15.3%) predominated, while Intraocular use (2.7%) and Ocular use (0.9%) were less common (**Table 2**). The distribution of dose form and route did not fully align. For example, Solution for injection and Injection accounted together for 10.4% of mapped dose forms for dexamethasone (vs 10% Tablet), yet Oral use covered 24.3% of the mapped routes.

**Table 1.**
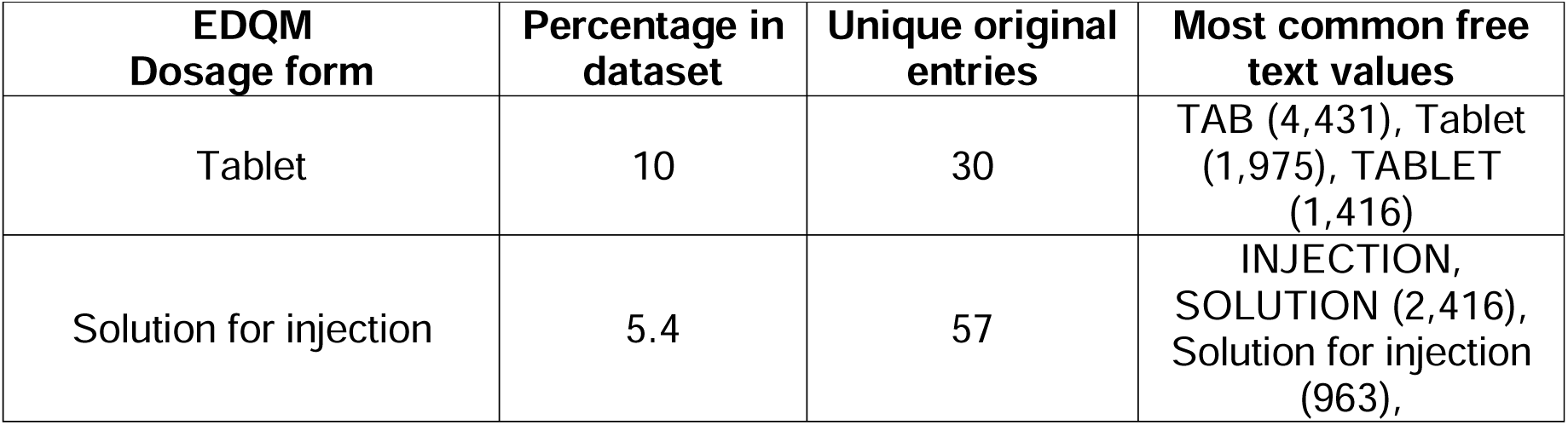
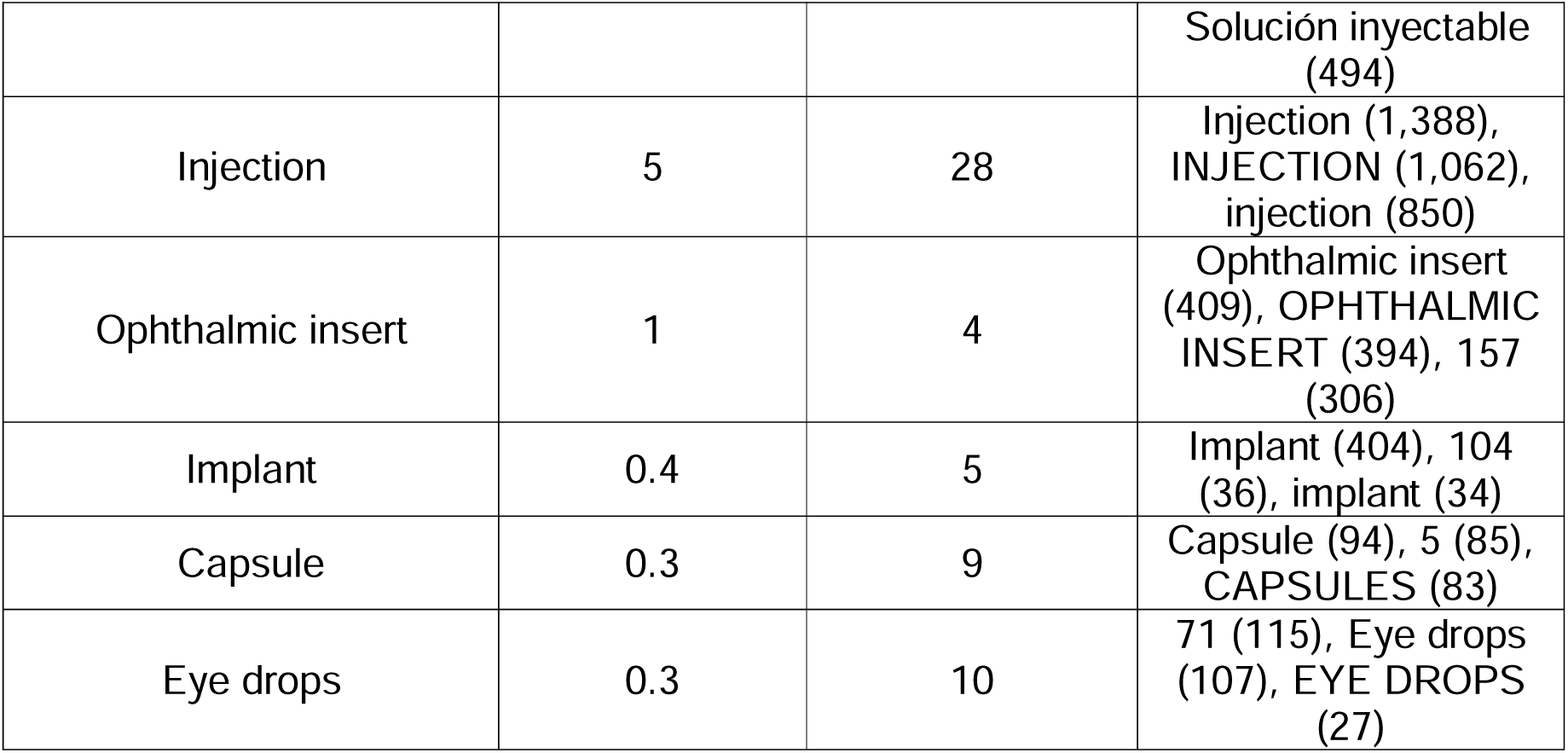
Mapped EDQM dosage forms for dexamethasone in VigiBase reports (January 1, 2001–December 31, 2024), with their percentage, unique original entries, and most common free text values. Original free-text and legacy terms were standardized to the closest corresponding EDQM dose form term

**Table 2.** Mapped EDQM routes of administration for dexamethasone in VigiBase reports (January 1, 2001– December 31, 2024), with their percentage, unique original entries, and most common free text values.

| EDQM Route of Administration | Percentage in dataset | Unique original entries | Most common free text values |
| --- | --- | --- | --- |
| Oral use | 24.3 | 3 | 048 (23,614), 050 (1,664), 48 (1,254) |
| Intravenous use | 15.3 | 7 | 042 (9,678), 050 (2,923), 041 (2,431) |
| Intraocular use | 2.7 | 3 | 031 (2,419), 050 (312), 31 (195) |
| Intramuscular use | 1.8 | 3 | 030 (1,309), 050 (489), 30 (135) |
| Ocular use | 0.9 | 3 | 047 (710), 47 (109), 050 (107) |

Mapping was further complicated by the heterogeneity of free text reporting. Dose forms appeared in multiple languages and included plural forms, abbreviations, codes, misspellings, and inconsistent formatting. For example, the EDQM term Tablet was derived from 30 different reported variants. Additionally, similar free text entries sometimes had to be mapped to different EDQM codes depending on the level of detail provided (e.g., a parenteral solution could be mapped to Solution for injection, Injection or Solution).

### 3.3 Disproportionality Analysis

Introducing administration site granularity revealed disproportionality patterns that differed from those observed in the substance-level reference analysis (Figure 6).

**Figure 6.**
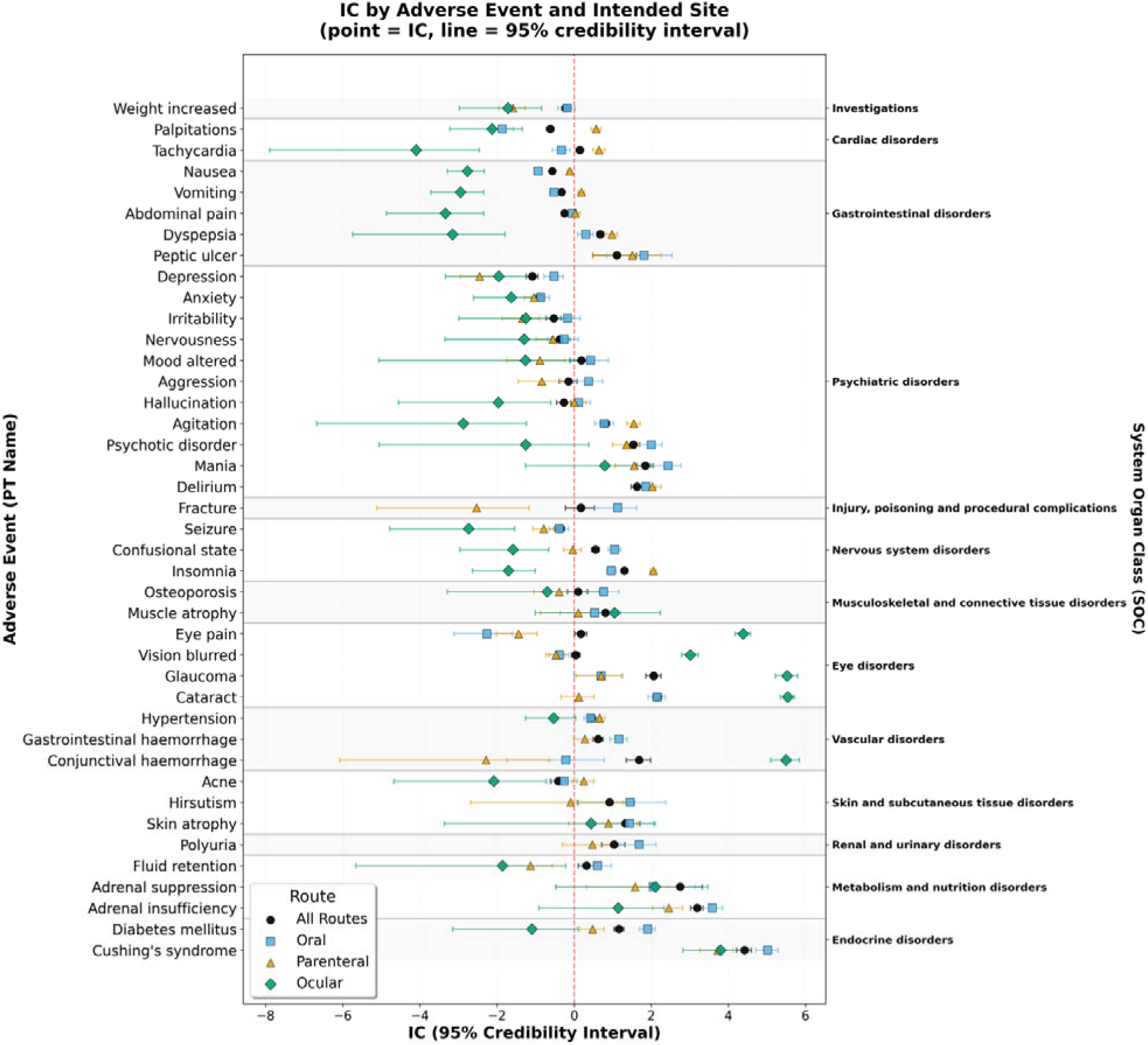
Information Component (IC) values with 95% credibility intervals for selected adverse reactions and dexamethasone, by administration site. Sites include reference (all sites combined), Ocular, Oral, and Parenteral use.

These site-specific patterns were most pronounced for less frequently represented administration sites and were generally consistent with known pharmacological mechanisms.

For Ocular use, ocular adverse reactions (e.g., *cataract, glaucoma, conjunctival haemorrhage, vision blurred, eye pain*) showed the strongest increases in disproportionality relative to the reference analysis. IC values for these events were close to zero for most other administration sites, with the exception of *cataract*, which was also disproportional for Oral use.

For systemic administration sites (i.e., Parenteral and Oral uses), site-specific IC values were generally closer to the reference values, reflecting the predominance of these exposures in the dataset. Psychiatric reactions such as *anxiety, agitation, mania,* and *hallucination* showed IC values that were similar to or higher than the reference for systemic administration sites, but lower for Ocular use. *Agitation* showed the highest IC for Parenteral use, whereas *depression* was highest for Oral use. In contrast, *Cushing’s syndrome*, *delirium* and *muscle atrophy* showed little variation across administration sites. For several events, Oral use IC values were slightly higher than the reference, whereas Ocular and Parenteral uses values were slightly lower and had wider intervals, suggesting that the substance-level disproportionality was driven largely by Oral use.

Some PTs showed negative reference IC values but positive site-specific IC values, indicating that signals may be diluted in substance-level analyses when reports from administration sites not plausibly associated with the event are included in the foreground. For example, *vomiting* and *palpitations* showed positive disproportionality only for Parenteral use (and not Oral use), and *fracture* showed positive disproportionality only for Oral use (and not Parenteral use).

Figure 7 and **Error! Reference source not found.** illustrate time-trends in IC for *cataract* and *palpitations* by administration site (Ocular, Oral, Parenteral use). For *cataract*, the Ocular use showed positive IC_025_ as early as 2012 (IC_025_ > 2), while the reference IC_025_ was still close to zero. *Palpitations* displayed a similar pattern: for Parenteral use, IC_025_ became positive from 2015 onward (IC_025_ >∼1), while the reference IC_025_ remained negative.

**Figure 7.**
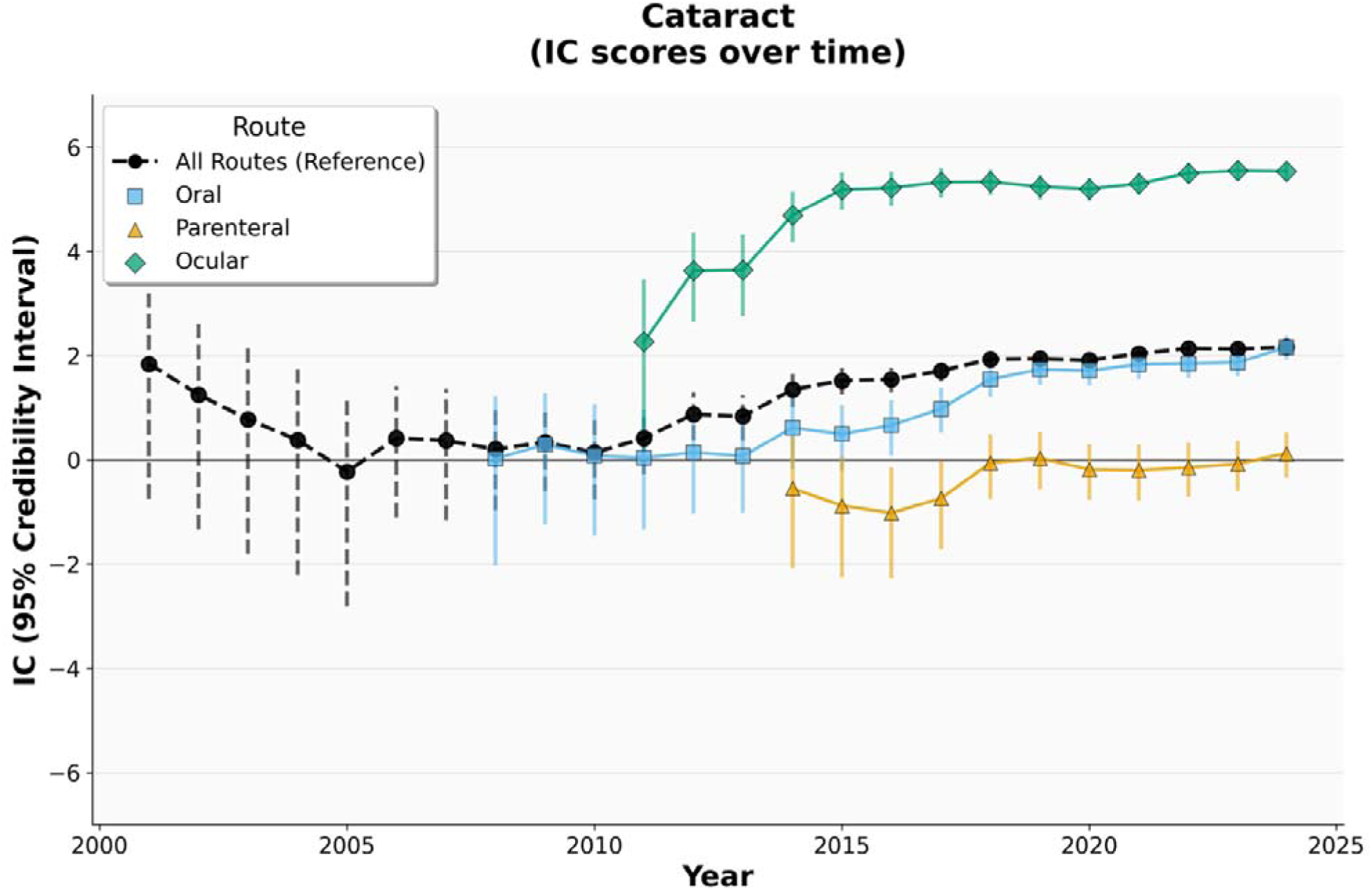
IC values over time (2001-2024) for the Dexamethasone - Cataract pair, by site (Ocular, Oral, Parenteral). Points represent IC estimates, and vertical lines indicate 95% credibility intervals. The black dashed line represents the reference IC (all sites combined). Intended site specific lines start at later time points due to absence of enough foreground data that could be mapped to those administration sites at earlier time points.

**Figure 8.**
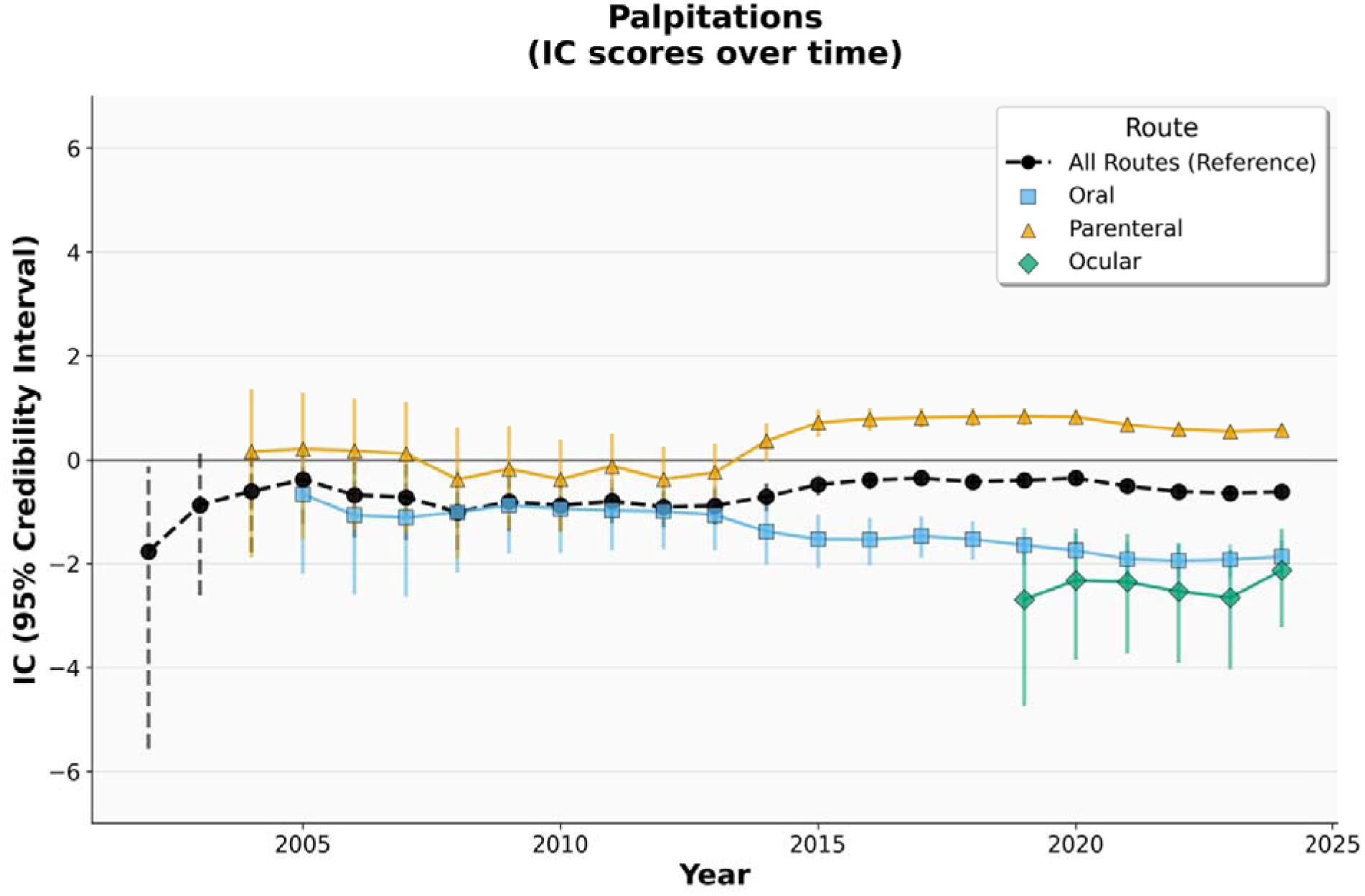
IC values over time (2001-2024) for the Dexamethasone - Palpitations pair, by administration site (Ocular, Oral, Parenteral use). Points represent IC estimates, and vertical lines indicate 95% credibility intervals. The black dashed line represents the reference IC (all sites combined). The reference IC starts in 2002 due to insufficient data regarding the drug-AE pair prior to that. Intended site specific lines start at later time points due to absence of enough foreground data that could be mapped to those administration sites at earlier time points.

## 4. Discussion

This study shows that by incorporating administration-site specificity into disproportionality analysis distinct safety patterns, biologically plausible but not apparent in conventional substance-level analyses, became visible. The present analysis should be interpreted as a proof of concept illustrating the potential of site-specific disproportionality analysis and, more in general, signal detection. In the dexamethasone case study, site-specific analyses showed stronger disproportionality for ocular events with Ocular use, and several psychiatric events showing stronger disproportionality for Oral or Parenteral use. In some instances, site-specific analyses also identified disproportionality earlier than the reference analysis, particularly for less common exposure patterns.

These findings illustrate the value of moving beyond the substance alone when evaluating safety signals. Substance-level analyses remain appropriate for many adverse reactions, but they can dilute site-specific associations by combining reports from formulations or routes for which the event is not expected. In our data, this was evident for events such as *Cataract* and *Palpitations*, where site-specific disproportionality emerged while the corresponding substance-level signal remained weak or absent. Use of more granular product information may therefore improve not only signal characterization but also the timeliness of detection.

The observed results were broadly consistent with a priori expectations, though with some insightful unexpected findings. As anticipated, ocular events (cataract, glaucoma, conjunctival haemorrhage, vision blurred, eye pain) showed markedly higher disproportionality for Ocular use than for systemic sites, confirming that site-specific analysis can recover well-established local safety signals. The expected predominance of psychiatric events for systemic sites was also confirmed, though with differentiation between Oral and Parenteral use that was not specified a priori: agitation showed the strongest association with Parenteral use, while depression was most pronounced for Oral use, a pattern that may reflect differences in treatment duration. Cushing’s syndrome, however, showed less site-specific variation than expected, with IC values that were broadly similar across sites, including ocular administration (coherent with case reports published in the literature, e.g., (24)). Among the events for which we had weaker a priori expectations, palpitations and vomiting proved notably site-specific, showing positive disproportionality only for Parenteral use, which would not have been apparent in the substance-level analysis. These cases illustrate the exploratory value of site-specific disproportionality even beyond hypothesis confirmation: they can surface patterns that refine clinical understanding of formulation-dependent risk.

At the same time, the study highlights the practical challenges of implementing such analyses in adverse event reports databases. Route of administration and dose form were frequently missing and highly heterogeneous, particularly because these fields were often recorded as multilingual free text with abbreviations, misspellings, and inconsistent levels of detail. Mapping therefore required substantial manual effort.

Some limitations should be considered beyond the normal caveats when dealing with adverse event reports and disproportionality analysis (25). First, often only one among route and dose form fields were compiled in the reports. When both were populated, in a few cases dose form and route were incoherent. These discrepancies reflect the incompleteness and inconsistency of reporting. Second, intended site is only an approximation of formulation granularity and distinct products with potentially different pharmacokinetic and safety properties could be grouped under the same site category such as ophthalmic suspension and intravitreal implant under Ocular use, or intravenous, intramuscular, and intra-articular use under Parenteral use. This could be mitigated by doing further analysis on product level.

Third, lack of report details reduced the number of exposed cases available for site-specific analyses. Because the width of the IC credibility interval depends on the number of observed reports, some site-specific associations may have been weakened or missed because of reduced statistical power rather than true absence of disproportionality.

An additional methodological consideration is the choice of background. In this study, site-specific foregrounds were compared against the full standardized database rather than against a background restricted to drugs reported for the same administration site. This approach preserves statistical power and avoids excessive subgrouping, but it does not account for adverse events that may be driven primarily by administration site or method rather than by the substance itself. The choice of background therefore influences the interpretation of site-specific disproportionality and merits further evaluation. A site-restricted background may be preferable for some research questions, particularly when the aim is to separate substance-related effects from reactions driven primarily by administration site or administration method. However, for this proof-of-concept analysis, we prioritized broader comparability and retention of statistical power.

The additional insights gained from this study depend on the completeness of intended-site information available in the reports, which as shown in Figure 4, appears to have slightly declined in recent years. This may be connected to the documented impact of the Covid-19 pandemic, during which overburdened health systems saw reduced reporting from healthcare practitioners (26), who are also those most likely to provide detailed information on administration route and site (27). Demonstrating that such information is meaningful rather than merely decorative is therefore timely and demonstrates the importance of implementing global standards such as ISO IDMP. By utilizing the global PhPID for reporting, the completeness and consistency of medicinal product information in newly submitted adverse event reports would improve, reducing the need for subjective coding choices, and make more granular pharmacovigilance analyses feasible at scale. For already collected reports, mapping will still be needed, and by using the global PhPID framework for dose form expression, a standardised mapping procedure could be applied. Mapping could be further improved using information from other report fields like indication for use and strength (which could univocally identify a specific formulation). These could also indicate divergence from the intended use. (e.g., off-label use, misuse, accidental exposure or medication error; for example, accidental eye exposure to paracetamol solution). Examining this interplay between intended and reported use may provide additional pharmacovigilance value, including insights into suboptimal medicine practices and could be further investigated in additional studies.

This study focused on administration site, but the same principle could be extended to other product dimensions based on the PhPIDs building blocks. Signal detection could for example be explored at the level of other dose form attributes such as release characteristics or administration method, or strength, and future work could assess how these levels of granularity complement one another (28). More systematic evaluations across additional drugs, events, and reference datasets will provide valuable insight into when granular analyses enhance signal detection and how they can best complement conventional substance-level screening.

Finally, the value of having global standards capturing aspects like administration site and strength, such as the PhPID, extends beyond disproportionality analysis. More detailed product characterization may support broader pharmacovigilance activities, including the identification of formulation-specific risks and the development of more targeted risk minimization measures, such as label changes or focused safety communications for particular products rather than for all products containing the same substance.

## 5. Conclusion

Incorporating administration site granularity, aligned with PhPID dose form standards, into disproportionality analysis can reveal site-specific safety concerns that may remain hidden or be detected later in substance-level analyses. These findings highlight the potential value of more granular global standards as an enabler of more precise signal detection and characterization in pharmacovigilance. Further evaluation across additional substances and events is needed, but these findings support the potential value of implementing global medicinal product identifiers such as PhPID in pharmacovigilance systems.

## Data Availability

For the purposes of this study, access to the data was granted following case-by-case review in accordance with those Conditions (IR12-2025). Subject to these conditions, data is available from the authors on reasonable request.

https://github.com/Uppsala-Monitoring-Centre/phpid_granularity_script

## Statements and Declarations

### Ethics approval

Not applicable. The data in VigiBase is de-identified and not considered personal data.

### Consent to participate

Not applicable. This study did not involve human participants.

### Consent to publish

Not applicable.

### Authors’ contributions

All authors participated in the conceptualization and design of the study. JFC and PVB drafted the manuscript. BRS, JFC and PVB conceptualized EDQM basic dose form mapping, JFC and PVB conceptualized route of administration EDQM mapping. PVB performed the data analysis. All authors contributed to the review of the manuscript. All authors have read and approved the final version.

### Availability of data and materials

The data that support the findings of this study are not publicly available. Access to data in VigiBase is subject to the requirements of the VigiBase Data Access Conditions. For the purposes of this study, access to the data was granted following case-by-case review in accordance with those Conditions (IR12-2025). Subject to these conditions, data is available from the authors on reasonable request. For further inquiries, please contact Uppsala Monitoring Centre via https://who-umc.org/contact-information

### Code availability

The code used for this study is publicly available in the current GitHub repository https://github.com/Uppsala-Monitoring-Centre/phpid_granularity_script but may be made available upon reasonable request. For further inquiries, please contact Uppsala Monitoring Centre via https://who-umc.org/contact-information/

## Acknowledgements

We would like to acknowledge Marilina Castellano (Uppsala Monitoring Centre-UMC) for her influence on the study design, Tomas Bergvall (UMC) for his guidance in applying EDQM standards during the standardization process and Jenny Klint (UMC) for reviewing the route and dose form mapping. Peter Hjelmström (UMC) reviewed and provided helpful feedback on the manuscript.

The authors are indebted to the members of the WHO Programme for International Drug Monitoring who contribute reports to VigiBase. However, the opinions and conclusions of this study are not necessarily those of the various member organisations nor of the WHO. MedDRA® trademark is registered by ICH. MedDRA® the Medical Dictionary for Regulatory Activities terminology is the international medical terminology developed under the auspices of the International Council for Harmonisation of Technical Requirements for Pharmaceuticals for Human Use (ICH).

## Footnotes

1 Sections G and B4 of ICH E2B(R3) and ICH E2B(R2).

2 such as European Directorate for the Quality of Medicines & HealthCare (EDQM) (13), U.S. Food and Drug Administration Structured Product Labeling resources (14), or ICH E2B(R2) codes (12).

