## Supplementary Material for "Leveraging global PhPID framework to enable more granular signal detection and characterization in VigiBase: a dexamethasone case study"

### S1: List of PT names analysed

 Abdominal pain

 Acne

 Adrenal insufficiency

 Adrenal suppression

 Aggression

 Agitation

 Anxiety

 Cataract

 Confusional state

 Conjunctival haemorrhage

 Cushing's syndrome

 Delirium

 Depression

 Diabetes mellitus

 Dyspepsia

 Eye pain

 Fluid retention

 Fracture

 Gastrointestinal haemorrhage

 Glaucoma

 Hallucination

 Hirsutism

 Hypertension

 Insomnia

 Irritability

 Mania

 Mood altered

 Muscle atrophy

 Nausea

 Nervousness

 Osteoporosis

 Palpitations

 Peptic ulcer

 Polyuria

 Psychotic disorder

 Seizure

 Skin atrophy

 Tachycardia

 Vision blurred

 Vomiting

 Weight increased

### S2: Intended site mapping

| **Route of administration EDQM** | **Dosage form EDQM** | **Administration site** |
| --- | --- | --- |
| Epidural use | Injection | Parenteral |
| Epidural use | Oral liquid | Parenteral |
| Epidural use | Solution for infusion | Parenteral |
| Epidural use | Solution for injection | Parenteral |
| Epidural use | Suspension for injection | Parenteral |
| Epidural use | Tablet | Parenteral |
| Epidural use | — | Parenteral |
| Intraarterial use | Injection | Parenteral |
| Intraarterial use | Solution for injection | Parenteral |
| Intraarterial use | — | Parenteral |
| Intraarticular use | Emulsion for injection | Parenteral |
| Intraarticular use | Emulsion for injection/infusion | Parenteral |
| Intraarticular use | Injection | Parenteral |
| Intraarticular use | Oral liquid | Parenteral |
| Intraarticular use | Oral solution | Parenteral |
| Intraarticular use | Solution for injection | Parenteral |
| Intraarticular use | — | Parenteral |
| Intracardiac use | Gel | Parenteral |
| Intracardiac use | Solution for injection | Parenteral |
| Intracardiac use | Tablet | Parenteral |
| Intracardiac use | — | Parenteral |
| Intracavernous use | Infusion | Parenteral |
| Intracavernous use | Injection | Parenteral |
| Intracavernous use | Solution for injection | Parenteral |
| Intracavernous use | — | Parenteral |
| Intracorneal use | — | Ocular |
| Intradermal use | Injection | Parenteral |
| Intradermal use | — | Parenteral |
| Intradiscal use | Injection | Parenteral |
| Intradiscal use | — | Parenteral |
| Intrahepatic | — | Parenteral |
| Intralesional use | Injection | Parenteral |
| Intramuscular use | Implant | Parenteral |
| Intramuscular use | Injection | Parenteral |
| Intramuscular use | Oral solution | Parenteral |
| Intramuscular use | Solution for infusion | Parenteral |
| Intramuscular use | Solution for injection | Parenteral |
| Intramuscular use | Solution for injection/infusion | Parenteral |
| Intramuscular use | Suspension for injection | Parenteral |
| Intramuscular use | Tablet | Parenteral |
| Intramuscular use | — | Parenteral |
| Intraocular use | Eye drops | Ocular |
| Intraocular use | Eye drops, solution | Ocular |
| Intraocular use | Eye drops, suspension | Ocular |
| Intraocular use | Eye ointment | Ocular |
| Intraocular use | Implant | Ocular |
| Intraocular use | Injection | Parenteral |
| Intraocular use | Ointment | Ocular |
| Intraocular use | Ophthalmic insert | Ocular |
| Intraocular use | Solution for injection | Parenteral |
| Intraocular use | Suspension for injection | Parenteral |
| Intraocular use | — | Ocular |
| Intraosseous use | Injection | Parenteral |
| Intraosseous use | Solution for injection | Parenteral |
| Intraosseous use | — | Parenteral |
| Intrapleural use | Injection | Parenteral |
| Intrapleural use | — | Parenteral |
| Intrathecal use | Infusion | Parenteral |
| Intrathecal use | Injection | Parenteral |
| Intrathecal use | Oral solution | Parenteral |
| Intrathecal use | Powder for injection | Parenteral |
| Intrathecal use | Powder for solution for injection | Parenteral |
| Intrathecal use | Solution for infusion | Parenteral |
| Intrathecal use | Solution for injection | Parenteral |
| Intrathecal use | Tablet | Parenteral |
| Intrathecal use | — | Parenteral |
| Intratumoral use | Injection | Parenteral |
| Intravenous use | Capsule | Parenteral |
| Intravenous use | Concentrate for solution for infusion | Parenteral |
| Intravenous use | Emulsion for injection | Parenteral |
| Intravenous use | Gel | Parenteral |
| Intravenous use | Implant | Parenteral |
| Intravenous use | Infusion | Parenteral |
| Intravenous use | Injection | Parenteral |
| Intravenous use | Nasal cream | Parenteral |
| Intravenous use | Oral liquid | Parenteral |
| Intravenous use | Oral solution | Parenteral |
| Intravenous use | Powder and solvent for solution for infusion | Parenteral |
| Intravenous use | Powder for concentrate for solution for infusion | Parenteral |
| Intravenous use | Powder for injection | Parenteral |
| Intravenous use | Powder for solution for infusion | Parenteral |
| Intravenous use | Powder for solution for injection | Parenteral |
| Intravenous use | Solution for infusion | Parenteral |
| Intravenous use | Solution for injection | Parenteral |
| Intravenous use | Solution for injection/infusion | Parenteral |
| Intravenous use | Solvent for parenteral use | Parenteral |
| Intravenous use | Suspension for injection | Parenteral |
| Intravenous use | Tablet | Parenteral |
| Intravenous use | — | Parenteral |
| Ocular use | Cream | Ocular |
| Ocular use | Ear drops | Ocular |
| Ocular use | Ear drops, solution | Ocular |
| Ocular use | Ear drops, suspension | Ocular |
| Ocular use | Ear/eye drops, solution | Ocular |
| Ocular use | Emulsion for injection | Ocular |
| Ocular use | Eye drops | Ocular |
| Ocular use | Eye drops, solution | Ocular |
| Ocular use | Eye drops, solution in single-dose container | Ocular |
| Ocular use | Eye drops, suspension | Ocular |
| Ocular use | Eye gel | Ocular |
| Ocular use | Eye lotion, solvent for reconstitution | Ocular |
| Ocular use | Eye ointment | Ocular |
| Ocular use | Implant | Ocular |
| Ocular use | Injection | Ocular |
| Ocular use | Ointment | Ocular |
| Ocular use | Ophthalmic insert | Ocular |
| Ocular use | Oral solution | Ocular |
| Ocular use | Solution for injection | Ocular |
| Ocular use | Solution for injection in pre-filled syringe | Ocular |
| Ocular use | Tablet | Ocular |
| Ocular use | — | Ocular |
| Oral use | Capsule | Oral |
| Oral use | Capsule, hard | Oral |
| Oral use | Coated tablet | Oral |
| Oral use | Concentrate for solution for infusion | Oral |
| Oral use | Cream | Oral |
| Oral use | Cutaneous foam | Oral |
| Oral use | Dental emulsion | Oral |
| Oral use | Ear cream | Oral |
| Oral use | Ear/eye drops, solution | Oral |
| Oral use | Effervescent tablet | Oral |
| Oral use | Eye drops | Oral |
| Oral use | Eye drops, solution in single-dose container | Oral |
| Oral use | Film-coated tablet | Oral |
| Oral use | Gel | Oral |
| Oral use | Implant | Oral |
| Oral use | Infusion | Oral |
| Oral use | Injection | Oral |
| Oral use | Nasal cream | Oral |
| Oral use | Ointment | Oral |
| Oral use | Oral drops | Oral |
| Oral use | Oral drops, emulsion | Oral |
| Oral use | Oral drops, solution | Oral |
| Oral use | Oral liquid | Oral |
| Oral use | Oral solution | Oral |
| Oral use | Oral suspension | Oral |
| Oral use | Powder for solution for infusion | Oral |
| Oral use | Powder for solution for injection | Oral |
| Oral use | Soluble tablet | Oral |
| Oral use | Solution for infusion | Oral |
| Oral use | Solution for injection | Oral |
| Oral use | Solution for injection/infusion | Oral |
| Oral use | Solvent for parenteral use | Oral |
| Oral use | Suspension for infusion | Oral |
| Oral use | Suspension for injection | Oral |
| Oral use | Syrup | Oral |
| Oral use | Tablet | Oral |
| Oral use | — | Oral |
| Parenteral | Injection | Parenteral |
| Parenteral | Solution for injection | Parenteral |
| Parenteral | Tablet | Parenteral |
| Parenteral | — | Parenteral |
| Periarticular use | Injection | Parenteral |
| Periarticular use | Solution for injection | Parenteral |
| Periarticular use | — | Parenteral |
| Perineural use | Injection | Parenteral |
| Perineural use | — | Parenteral |
| Retrobulbar use | Injection | Parenteral |
| Retrobulbar use | — | Ocular |
| Subconjunctival use | Eye drops | Ocular |
| Subconjunctival use | Injection | Parenteral |
| Subconjunctival use | Solution for injection | Parenteral |
| Subconjunctival use | — | Ocular |
| Subcutaneous use | Capsule | Parenteral |
| Subcutaneous use | Gel | Parenteral |
| Subcutaneous use | Injection | Parenteral |
| Subcutaneous use | Oral liquid | Parenteral |
| Subcutaneous use | Solution for injection | Parenteral |
| Subcutaneous use | Suspension for infusion | Parenteral |
| Subcutaneous use | Tablet | Parenteral |
| Subcutaneous use | — | Parenteral |
| — | Emulsion for injection | Parenteral |
| — | Emulsion for injection/infusion | Parenteral |
| — | Eye drops | Ocular |
| — | Eye drops, solution | Ocular |
| — | Eye drops, solution in single-dose container | Ocular |
| — | Eye drops, suspension | Ocular |
| — | Eye gel | Ocular |
| — | Eye ointment | Ocular |
| — | Infusion | Parenteral |
| — | Injection | Parenteral |
| — | Ophthalmic insert | Ocular |
| — | Oral liquid | Oral |
| — | Oral solution | Oral |
| — | Soluble tablet | Oral |
| — | Solution for infusion | Parenteral |
| — | Solution for injection | Parenteral |
| — | Solution for injection/infusion | Parenteral |
| — | Suspension for injection | Parenteral |
| — | Tablet | Oral |
